# COVID-19 Underreporting and its Impact on Vaccination Strategies

**DOI:** 10.1101/2021.03.11.21253404

**Authors:** Vinicius Albani, Jennifer Loria, Eduardo Massad, Jorge P. Zubelli

**Affiliations:** Department of Mathematics, Federal University of Santa Catarina, Florianopolis, Brazil; Instituto de Matemática Pura e Aplicada, Rio de Janeiro, Brazil; Universidad de Costa Rica, San Jose, Costa Rica; School of Medicine, University of Sao Paulo and LIM01-HCFMUSP, São Paulo, Brazil; School of Applied Mathematics, Fundação Getúlio Vargas, Rio de Janeiro, Brazil; Mathematics Department, Khalifa University, Abu Dhabi, UAE

## Abstract

We present a novel methodology for the stable rate estimation of hospitalization and death related to the Corona Virus Disease 2019 (COVID-19) using publicly available reports from various distinct communities. These rates are then used to estimate underreported infections on the corresponding areas by making use of reported daily hospitalizations and deaths. The impact of underreporting infections on vaccination strategies is estimated under different disease-transmission scenarios using a Susceptible-Exposed-Infective-Removed-like (SEIR) epidemiological model.

**One sentence Summary:** Using a novel methodology, we estimate COVID-19 underreporting from public data, quantifying its impact on vaccination.

---

Underreporting cases of infectious diseases poses a major challenge in the analysis of their epidemiological characteristics and dynamical aspects. Without accurate numerical estimates it is difficult to precisely quantify the proportions of severe and critical cases, as well as the mortality rate (*1*). Such estimates can be provided, e.g., by testing the presence of the virus. However, during an ongoing epidemic, such tests’ implementation is a daunting task. Thus, this work presents a methodology to estimate underreported infections based on approximations of the stable rates of hospitalization and death. In order to find such rates, we seek specific time periods when the daily rate of testing is sufficiently large with respect to the population size, and the number of positive tests is small enough. During such periods we evaluate daily empirical rates of hospitalization and death, looking for those whose rates fluctuate around some mean value. This is performed by means of an accurate data analysis producing different statistical indicators leading to the necessary correction. A schematic representation that summarizes the proposed methodology can be found in Figure 1.

**Figure 1:**
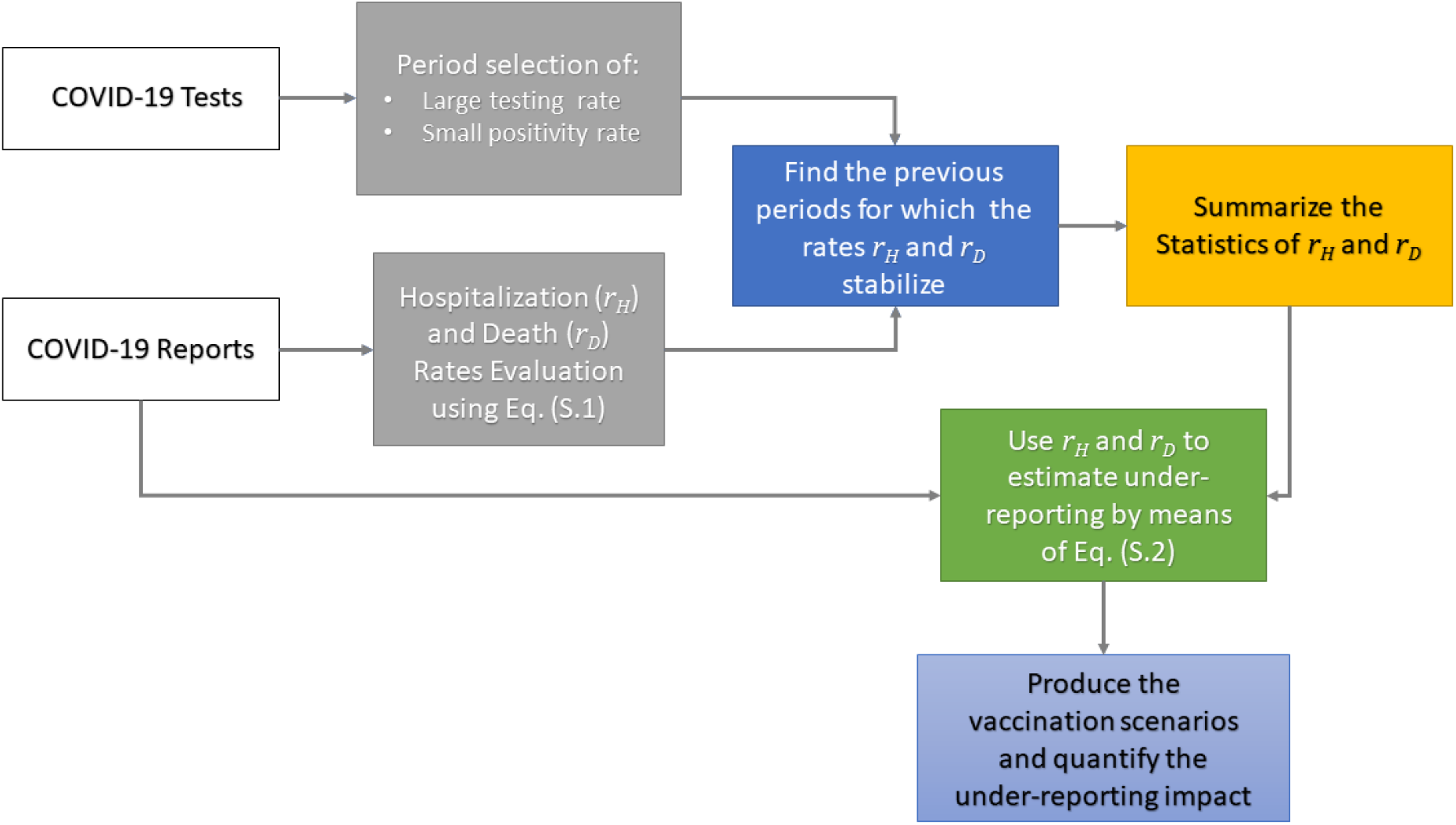
Methodological workflow for the underreporting quantification.

We use time series of seven-day moving averaged reports from Chicago and New York City (NYC), in the US, the province of Buenos Aires (BA), in Argentina, and Mexico City (MC), in Mexico.

Since COVID-19 severity strongly depends on age and gender (*2*–*6*), we evaluate the above-mentioned rates accounting for demography to improve the estimation accuracy of the number of infections. The latter will be called *corrections*. These corrections are evaluated using the empirical rates of hospitalization and death as follows: For an observed rate of hospitalization or death, and a given day in the time series, we evaluate the corresponding infection number. For example, if for this day the reported hospitalization rate is one and the projected rate is one half, then, the correction is twice the reported infections.

As an important byproduct, we evaluate the impact of underreporting in the designing of vaccination strategies because the larger the number of unaccounted infections, the larger the chances of vaccinating an already immune individual. This can restrict the capability of vaccination in reducing hospitalizations and deaths, as simulated scenarios using a Susceptible-Exposed-Infective-Removed-like (SEIR-like) model (*7*) show.

In order to estimate underreported infections, the formulas in Eq. (S.2) are used, considering the daily cases of COVID-19. The graphical comparison between the observed and corrected numbers of infections for Chicago can be found in Figure 2. Table 1 presents the corrected and observed accumulated numbers of COVID-19 infections in Chicago, during the period 01Mar-2020 to 23-Dec-2020. In order to observe the effect of corrections, we divided the period 01-Mar-2020 to 23-Dec-2020 into three periods, namely, 01-Mar-2020 to 31-July-2020, 01Aug-2020 to 05-Oct-2020, and 06-Oct-2020 to 23-Dec-2020. Additional results considering the data from other places can be found in the Supplementary Materials, as well as the details on the implementation of the techniques used in this work.

**Table 1:** Accumulated numbers of corrected and reported infections in Chicago from 01-Mar-2020 to 23-Dec-2020. Corrections use the median values and the 90% CI values from Table S.1.

| Citywide Correction |  |  |  |
| --- | --- | --- | --- |
| Period | By Hopitalization Rate | By Death Rate | Observed |
| 01-Mar to 31-July | 169,126 (149,945–222,446) | 290,123 (186,707–423,991) | 61,905 |
| 01-Aug to 05-Oct | 21,772 (21,157–27,321) | 22,607 (21,134–30,861) | 20,790 |
| 06-Oct to 23-Dec | 113,992 (113,992–113,994) | 127,476 (115,017–167,933) | 113,693 |
| 01-Mar to 23-Dec | 304,890 (285,095–363,762) | 440,207 (322,859–622,785) | 196,388 |
| Correction By Gender |  |  |  |
| 01-Mar to 31-July | 170,244 (143,304–223,130) | 297,418 (197,599–453,493) | 61,905 |
| 01-Aug to 05-Oct | 21,994 (21,049–27,231) | 23,616 (21,085–33,237) | 20,790 |
| 06-Oct to 23-Dec | 113,561 (113,561–113,712) | 129,178 (115,451–178,659) | 113,693 |
| 01-Mar to 23-Dec | 305,798 (277,914–364,073) | 450,212 (334,135–665,389) | 196,388 |
| Correction By Age Range |  |  |  |
| 01-Mar to 31-July | 141,280 (97,351–254,365) | 168,202 (80,124–264,252) | 61,905 |
| 01-Aug to 05-Oct | 23,998 (21,209–40,851) | 25,180 (21,146–31,802) | 20,790 |
| 06-Oct to 23-Dec | 114,188 (113,979–132,578) | 118,656 (113,986–146,240) | 113,693 |
| 01-Mar to 23-Dec | 279,466 (232,539–427,795) | 312,038 (215,257–442,295) | 196,388 |

**Figure 2:**
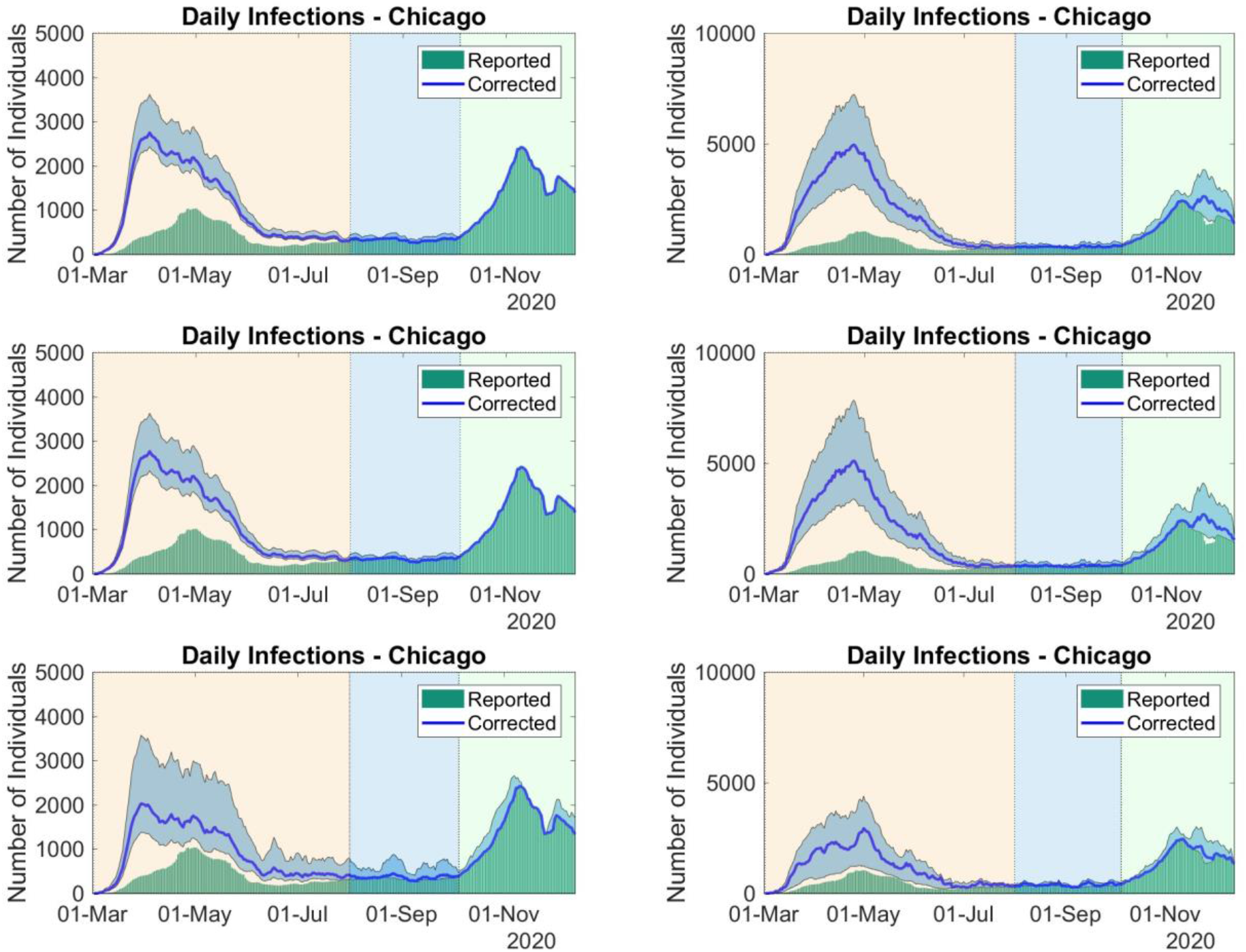
Corrected and reported series of daily infections in Chicago from 01-Mar-2020 to 23-Dec-2020, using the rates of hospitalization (left column) and death (right column) from Table S.1. First row uses the daily reports, the second uses daily reports by gender, and the third one uses daily reports by age range. The filled envelopes are 90% confidence intervals (CIs).

Corrections using hospitalization rates present smaller values than the ones obtained with death rates. This can be explained by the considerably larger values of the death rate in hospital observed during the outbreaks of March to May and of October to November. The estimated numbers for 01-Mar-2020 to 31-July-2020 are larger than the ones estimated for other periods, indicating that underreport can be more likely in the beginning of the pandemic. Corrections suggest that, for 01-Mar-2020 to 31-July-2020, the number of infections can be 32% to 632% larger. For 01-Mar-2020 to 23-Dec-2020, COVID-19 infections can be 10% to 238% larger. Thus, from 8% to 25% of the population of Chicago could have being infected in the study period, instead of the observed proportion of 7.3%.

The datasets from NYC do not have daily reports by age range or gender. We considered two different periods to estimate the stable rates of hospitalization and deaths and corrected infections can be found in Table S.3, representing 7.5% to 30% of the NYC population, instead of the observed proportion of 4.41%.

For BA, unfortunately, during the period of study the percentage of positive tests was mostly above 10%, making difficult the empirical analysis. However, we consider the period when the positive rate was below 20%. Table S.4 presents the estimated rates of hospitalization and death. Death rates for individuals younger than 60 years old are like the corresponding rates observed in Chicago. However, for older individuals in BA, the death rates are considerably larger. Corrections from Table S.5 suggest infection numbers varying from 3.4% to 303% larger than the notified cases, representing 4.7% to 18% of the estimated BA population for 2020, instead of the reported 4.53%.

For MC we could not identify a period when the rates of death or hospitalization stabilized around mean values. Thus, we used the rates estimated for Chicago to provide corrections. Using the death rates by age-range from Chicago seems to be the more accurate way to estimate underreported cases in other places, since the data from Chicago satisfied the hypotheses made to find stable rates. Corrections are 44% to 681% larger than the observed cases, representing 5.5% to 30% of the estimated population of MC for 2015.

Let us now turn to the impact of underreporting on the capacity of vaccination strategies in reducing hospitalizations and deaths. We consider two different scenarios. In the first one, simulations are performed under contained spread. In the second one, vaccination is implemented under increasing transmission. The parameters used in these examples are estimated using reports from Chicago and NYC (*8*).

In both cases we assume that the proportion of the population in the recovered, exposed or in some infective compartment in the model in Eqs. (S.3)-(S.11), ranges from 5% to 30%. Moreover, only the amount of 5% is observed in all cases. This means that the probability of vaccinating someone that has already had contact with the virus is proportional to the percentage of the population distributed in the exposed, non-hospitalized and infective, and recovered compartments that were not included in the reports. Thus, in our simulations if 5% of the population was infected, then 100% of the vaccinated individuals were susceptible, whereas, if 30% of the population was infected, then only 73.4% of the vaccinated individuals were susceptible. We also assume that the vaccine is 90% effective, and 0.5% of the population is vaccinated every day, for 150 days. The hospitalization rate also decreased proportionally to the number of underreports.

Under contained spread, the transmission parameter amongst mildly infective individuals is set to *β*_*M*_ = 0.23. Under uncontained transmission, the parameter *β*_*M*_ is set to 0.44. The resulting accumulated numbers during the vaccination strategy, in both situations, can be found in Table 2.

**Table 2:** Accumulated numbers of recovered, vaccinated, hospitalized, and deceased individuals after a random mass vaccination strategy of 150 days, when the proportion of individuals that has already had contact with the virus ranges from 5% to 30% of the population, whereas reports represent only 5%.

| Contained Spread |  |  |  |  |  |  |
| --- | --- | --- | --- | --- | --- | --- |
| Proportion | 5% | 10% | 15 % | 20% | 25% | 30% |
| Initial Recovered | 117,704 | 237,730 | 357,756 | 477,782 | 597,808 | 717,834 |
| Total Recovered | 195,683 | 377,439 | 542,234 | 696,487 | 842,935 | 983,440 |
| Total Vaccinated | 1,808,276 | 1,537,035 | 1,446,621 | 1,356,207 | 1,265,794 | 1,175,380 |
| Hospitalizations | 3,192 | 4,847 | 5,993 | 6,625 | 6,892 | 6,893 |
| Deaths | 82 | 120 | 147 | 162 | 167 | 167 |
| Uncontained Spread |  |  |  |  |  |  |
| Proportion | 5% | 10% | 15 % | 20% | 25% | 30% |
| Initial Recoverd | 117,181 | 236,675 | 356,168 | 475,661 | 595,154 | 714,648 |
| Total Recovered | 428,580 | 697,122 | 866,907 | 1,006,654 | 1,130,436 | 1,244,991 |
| Total Vaccinated | 1,800,249 | 1,530,212 | 1,440,200 | 1,350,187 | 1,260,175 | 1,170,162 |
| Hospitalizations | 13,107 | 16,418 | 17,063 | 16,554 | 15,500 | 14,185 |
| Deaths | 200 | 250 | 259 | 251 | 235 | 215 |

The assumed size of this hypothetical population is of 2,693,976 individuals. In Table 2, the numbers in the row *Total Vaccinated* correspond to the vaccinated individuals that were in the susceptible compartment. As the underreported infections increase, the number of effectively vaccinated individuals decreases. The recovered individuals are considered permanently immune. The capacity of vaccination in reducing hospitalizations and deaths is hampered due to underreporting, both under contained and uncontained disease spread. However, if the disease transmission is not under control, then, as underreport increases, the number of hospitalizations and deaths can decrease, indicating the achievement of herd immunity. Therefore, estimating underreporting helps to quantify and explain possible limitations of vaccination strategies.

This work proposes possible ways to estimate underreported COVID-19 infections, based on daily reported of cases, hospitalizations, and deaths, considering demography. The proposed methodology of correction is then applied to data from Chicago, NYC, BA, and MC. Moreover, it estimates the potential impact of underreporting in vaccination strategies by using an SEIR-like model with parameters estimated from real data.

Estimating underreporting in an ongoing epidemic is a hard task, and only a seroprevalence study can address this task appropriately. However, if we can estimate the stable rates of hospitalization and death related to the disease, then we can use reports to estimate the correct number of infections. The major difficulty of this approach is to identify the period when these rates can be observed or approximated. Firstly, we assume that the number of tests performed daily must be sufficiently large, then the number of positive tests must be sufficiently small. Setting up this is subtle, and we must compare the data from different places. For Chicago and NYC, we set that the rate of positive tests must be below 10% and for BA, it was 20%, since we identified, in the corresponding periods, a stabilization of the rates around mean values. For MC, we could not find such period.

For Chicago, NYC, and MC, during the period of study, corrections suggest that the number of infected individuals could reach 30% of the population of these places, which represents, in some cases, more than six times the reported numbers. Such estimates must be considered when evaluating the aftermath of vaccination strategies, since underreporting, as illustrated by numerical examples, can reduce the impact of vaccination in reducing mortality and hospitalization rates. Estimating underreports can be useful, for example, to adjust the daily numbers of given vaccines in order to reach the target of reducing the numbers of infections, hospitalizations, and deaths.

Using age-dependent death rates seems to be a reliable way of estimating underreporting, since such rates can be used even if the age pattern of the infected population changes during the epidemic. Thus, we expect that the more demographic information we incorporate into the death rates, the more reliable are the corrections.

In summary, using the methodology described in Figure 1 and employing a judiciously chosen data analysis implementation, we estimate COVID-19 underreporting from publicly available data. This leads to a powerful way of quantifying underreporting impact on the efficacy of vaccination strategies.

## Data Availability

All the data used in this study came from public sources.

## Authors contributions

VA, EM and JZ proposed the mathematical model. VA and JL performed numerical simulations. VA and JL analyzed the data. All the authors contributed to the writing of the article. EM and JZ critically revised the manuscript. Competing interests: All authors declare no competing interests.

## Data and materials availability

The data that support the findings of this study are available from the following publicly sources: data.cityofchicago.org (Chicago), www1.nyc.gov (NYC), datos.cdmx.gob.mx (BA), and datos.cdmx.gob.mx (MC). The numerical scripts used to generate corrections and to simulated scenarios can be found in the GitHub repository github.com/JennySorio/Under Reporting.

## Supplementary Materials

Materials and Methods

Underreport Estimation and Stable Rates of Hospitalization and Death in Other Locations

Figs. S.1 to S.7

Tables S.1 to S.6

References (*9-16*)

